# Application of a 27-protein candidate cardiovascular surrogate endpoint to track risk ascendancy and resolution in COVID-19

**DOI:** 10.1101/2021.01.28.21250129

**Authors:** Clare Paterson, Yolanda Hagar, Michael A. Hinterberg, Alexander W. Charney, Diane M. Del Valle, Michael R. Filbin, Sacha Gnjatic, Jason D. Goldman, Nir Hacohen, James R. Heath, Rainer Hillenbrand, Lori L. Jennings, Seunghee Kim-Schulze, Andrew T. Magis, Miriam Merad, Konstantinos Mouskas, Nicole W. Simons, Stephen A. Williams

**Affiliations:** SomaLogic Inc. Boulder, CO, USA; Icahn School of Medicine at Mount Sinai, New York, NY, USA; Department of Emergency Medicine, Massachusetts General Hospital, Boston, MA, USA; Department of Emergency Medicine, Harvard Medical School, Boston, MA, USA; Broad Institute of Massachusetts Institute of Technology and Harvard, Cambridge, MA, USA; Swedish Center for Research and Innovation, Swedish Medical Center, Seattle, WA, USA; Providence St. Joseph Health, Renton, WA, USA; Division of Allergy & Infectious Diseases, University of Washington, Seattle, WA, USA; Massachusetts General Hospital Cancer Center, Department of Medicine, Massachusetts General Hospital, Boston, MA, USA; Department of Medicine, Harvard Medical School, Boston, MA, USA; Institute for Systems Biology, Seattle, WA, USA; Department of Bioengineering, University of Washington, Seattle, WA, USA; Novartis Institutes for BioMedical Research, Basel, Switzerland; Novartis Institutes for BioMedical Research, Cambridge, MA, USA

**Keywords:** COVID-19, cardiovascular risk, proteomics

## Abstract

**Background:** There is an urgent need for tools allowing the early prognosis and subsequent monitoring of individuals with heterogeneous COVID-19 disease trajectories. Pre-existing cardiovascular (CV) disease is a leading risk factor for COVID-19 susceptibility and poor outcomes, and cardiac involvement is prevalent in COVID-19 patients both during the acute phase as well as in convalescence. The utility of traditional CV risk biomarkers in mild COVID-19 disease or across disease course is poorly understood. We sought to determine if a previously validated 27-protein predictor of CV outcomes served a purpose in COVID-19.

**Methods:** The 27-protein test of residual CV (RCV) risk was applied without modification to n=860 plasma samples from hospitalized and non-hospitalized SARS-CoV-2 infected individuals at disease presentation from three independent cohorts to predict COVID-19 severity and mortality. The same test was applied to an additional n=991 longitudinal samples to assess sensitivity to change in CV risk throughout the course of infection into convalescence.

**Results:** In each independent cohort, RCV predictions were significantly related to maximal subsequent COVID-19 severity and to mortality. At the baseline blood draw, the mean protein-predicted likelihood of an event in subjects who died during the study period ranged from 88-99% while it ranged from 8-36% in subjects who were not admitted to hospital. Additionally, the test outperformed existing risk predictors based on commonly used laboratory chemistry values or presence of comorbidities. Application of the RCV test to sequential samples showed dramatic increases in risk during the first few days of infection followed by risk reduction in the survivors; a period of catastrophically high cardiovascular risk (above 50%) typically lasted 8-12 days and had not resolved to normal levels in most people within that timescale.

**Conclusions:** The finding that a 27-protein candidate CV surrogate endpoint developed in multi-morbid patients prior to the pandemic is both prognostic and acutely sensitive to the adverse effects of COVID-19 suggests that this disease activates the same biologic risk-related mechanisms. The test may be useful for monitoring recovery and drug response.

## INTRODUCTION

The novel coronavirus disease 2019 (COVID-19) caused by the SARS-CoV-2 virus has become a global public healthcare and socioeconomic emergency, with over 65 million confirmed cases contributing to over 1.5 million deaths worldwide as of mid-December 2020.^1^ While risk factors for hospitalization and severe outcomes of COVID-19, including age, male sex, and the presence of comorbid chronic conditions affecting pulmonary, CV and metabolic systems have been identified,^2-4^ the significant heterogeneity of clinical phenotypes amongst COVID-19 patients poses a burden in clinical care decision making and healthcare resource allocation. Therefore, prognostic tools that can more precisely identify individuals who are likely to encounter morbid outcomes serve the utility to inform healthcare management, enabling earlier and more personalized care. In addition, as emerging evidence suggests prevalent long-term pathological sequalae in COVID-19 survivors independent of disease severity,^5^ understanding the biological determinants, identifying predictors and monitoring the resolution of prolonged adverse health outcomes is essential.

While the most obvious clinical manifestation in hospitalized COVID-19 patients is viral pneumonia, likely a consequence of an overactive host immune response, many individuals display extrapulmonary manifestations including cardiac-related events.^2,6^ Multiple studies have reported that both the susceptibility to, and the severity of outcome to COVID-19 are significantly associated with cardiovascular disease. Pre-existing cardiovascular disease not only increases an individual’s risk for death and severe course of illness,^7-9^ but SARS-CoV-2 infection itself induces cardiac damage in a significant proportion of hospitalized patients. Cardiac injury in COVID-19 patients is significantly associated with higher risk for in-hospital mortality and occurs in a sizeable proportion of patients without pre-existing cardiovascular disease.^10-12^ Elevations in traditional cardiac biomarkers such as high-sensitivity cardiac Troponin and N-terminal pro-BNP have demonstrated prognostic ability to predict severity of hospitalized COVID-19 patient outcomes^2,3^; however, the clinical utility of these findings has recently come under debate ^13^ and the validity of these biomarkers in cases of milder disease course is unknown. Together this suggests that a more comprehensive blood-based biomarker of underlying sub-clinical cardiac dysfunction at COVID-19 presentation may serve value as a candidate surrogate endpoint for COVID-19 disease severity and mortality as well as enabling monitoring COVID-19 related CV risk across the course of illness into convalescence.

We have recently developed and validated a 27 protein-only test highly predictive of residual future CV-event risk (four-year likelihood of myocardial infarction, stroke, hospitalization for heart failure or all-cause death) in a multiple cohort study of 18,165 higher risk individuals with relevance to COVID-19 outcome severity, including those with underlying CV complications, the elderly and diabetics.^14^

In the current study we tested the ability of this validated residual cardiovascular disease (RCV) test to predict mortality and stratify severity risk groups in three independent cohorts of COVID-19 infected individuals with the aim of evaluating it as a novel prognostic tool for COVID-19 related outcomes with applicability beyond hospitalized patients. Additionally, we assessed in a total of 1,851 longitudinal samples obtained from all three cohorts the utility of this test to characterize the ascendancy and resolution of risk across the course of SARS-CoV-2 infection into convalescence in order to determine the value of the test beyond the acute phase and highlight its potential to serve purpose as a monitoring tool for post-COVID-19 adverse sequelae.

## METHODS

### COVID-19 patient cohorts

Baseline and longitudinal patient samples utilized in the current study were obtained from patients with confirmation of SARS-CoV-2 positivity at time of study enrollment via PCR of nasopharyngeal or oropharyngeal swabs. Summary demographic characteristics of each cohort are shown in Table 1.

**Table 1.** Baseline demographic characteristics of COVID-19 patients in each cohort. MSSM, Mt Sinai School of Medicine; MGH, Massachusetts General Hospital; ISB, Institute for Systems Biology.

MAIN FIGURES
| Variable |  | MSSM (n=458) | MGH (n=298) | ISB (n=104) |
| --- | --- | --- | --- | --- |
| **Mortality**<br>n (%) | **Yes** | 93 (20) | 41 (14) | 8 (8) |
|  | **No** | 365 (80) | 257 (86) | 96 (92) |
| **Sex**<br>n (%) | **Female** | 180 (39) | 142 (48) | 62 (60) |
|  | **Male** | 278 (61) | 156 (52) | 42 (40) |
| **Ethnicity**<br>n (%) | **Hispanic/Latino** | 186 (41) | 161 (54) | 12 (12) |
|  | **Not Hispanic/Latino** | 264 (58) | 137 (46) | 88 (85) |
|  | **Unknown** | 8 (2) | 0 (0) | 4 (4) |
| **Race**<br>n (%) | **Asian** | 31 (7) | 11 (4) | 15 (14) |
|  | **Black or African American** | 109 (24) | 28 (9) | 7 (7) |
|  | **White** | 165 (36) | 151 (51) | 63 (61) |
|  | **Other/Unknown** | 153 (33) | 108 (36) | 19 (18) |
| **Age (years)** | **Mean (SD)** | 63.3 (16.1) | 59.6 (18.69) | 57.1 (19.2) |
|  | **Range** | (20, 90) | (25, 95) | (18, 89) |

#### Cohort 1: Mount Sinai Health System

Citrate plasma specimens were collected from hospitalized individuals presenting with suspicion of COVID-19 to Mount Sinai Hospital Center between dates 4/4/2020 to 8/30/2020. This work was reviewed and approved by the Institutional Review Board of the Human Research Protection Program at the Icahn School of Medicine at Mount Sinai. Details of this cohort have been previously described.^15^ For the current study, baseline citrate plasma specimens were available from 458 patients near to hospital admission. An additional 491 plasma samples were collected from 262 of the original 458 patients longitudinally over the course of infection spanning between days 1-102 of hospitalization.

#### Cohort 2: Massachusetts General Hospital

EDTA plasma samples were collected from individuals presenting to the Emergency Department of an urban, academic hospital in Boston with symptoms of acute respiratory distress and subsequently confirmed SARS-CoV-2 positive. Samples were collected between 3/24/2020 to 4/30/2020 following Partners Human Research Committee Institutional Review Board approval. Details of this cohort have been previously published.^16^ In brief, EDTA plasma samples were collected from 298 patients at day 0 (time of presentation to the emergency department). In a subset of patients still hospitalized, EDTA plasma samples were obtained at 3 and 7 days after the initial blood draw to allow for longitudinal analysis of an additional 335 samples from 219 of the original 298 patients.

#### Cohort 3: Institute for Systems Biology

EDTA plasma samples were obtained from confirmed COVID-19 patients spanning all levels of disease severities including home (mobile phlebotomy), clinic visit, hospitalized non-ICU patients, hospitalized ICU patients between 3/28/20 to 9/3/20.^17^ This study was approved by the Providence Institutional Review Board. For the current study, baseline plasma samples (collected shortly after initial clinical diagnosis) from 104 patients were available for proteomic analysis. Additional samples were collected during the acute phase of infection 3-14 days following baseline sample, as well as in convalescence (14-60 days following baseline) to allow for longitudinal analysis of an additional 165 samples from 102 of the original 104 patients.

### COVID-19 clinical outcomes

Patient clinical data was determined through extensive retrospective electronic health record and chart review. Two clinical outcomes of COVID-19 were investigated in the current study, death and maximum disease severity grade.

#### Death

Mortality rates of 20%, 14% and 8% were observed in cohorts 1, 2 and 3, respectively. The timeframes for assessment of death differed in each cohort based on the individual study design. Cohort 1: in-hospital death, cohort 2: within 28 days of baseline blood draw, and cohort 3: within follow up period, up to several months.

#### Maximum COVID-19 disease severity

The grading for maximum COVID-19 disease severity differed in each cohort based on the individual study design; however, there was a large overlap in the clinical parameters used to stratify patient severity. Each Cohort severity score grading definitions are described in detail in Supplementary Table 2. In brief, Cohort 1 maximum disease severity classification was based upon maximum level of oxygen support patient received, or death due to COVID-19, during hospitalization. Cohorts 2 and 3 maximum disease classification was based upon the maximum World Health Organization (WHO) Ordinal Scale of COVID-19 Disease Severity score throughout the study period.^18^ Due to evolving WHO guidelines throughout the study collection period, each of the cohorts followed slightly differing severity scoring criteria predominantly driven by the later inclusion further stratification based on requirement of additional organ support.

**Table 2.** The results and predictive metrics for the RCV model to predict both death and maximum severity endpoints across all three cohorts of COVID-19 patients. MSSM, Mt Sinai School of Medicine; MGH, Massachusetts General Hospital; ISB, Institute for Systems Biology. ^*^ The ISB results are based on 6 cases of death in the training data, and two cases of death in the test data.

| COVID-19 Endpoint | Metric |  | MSSM | MGH | ISB |
| --- | --- | --- | --- | --- | --- |
| Death (logistic regression) | Model estimate (p-value) | Percent increase in risk based for increase in RCV score | 12,296% (< 0.01) | 388% (< 0.01) | 116% (< 0.01) |
|  | Cross-validation model metrics (training/test) | AUC | 0.76/0.82 | 0.85/0.79 | 0.90/0.65* |
|  |  | Sensitivity | 0.66/0.73 | 0.84/0.75 | 0.89/0.45* |
|  |  | Specificity | 0.80/0.89 | 0.75/0.78 | 0.83/1.0* |
| Maximum severity (ordinal regression) | Model estimate (p-value) | Increase in risk with percentage point increase in RCV scores | 20.2% (< 0.01) | 1.11% (< 0.01) | 6.6% (< 0.01) |

### Plasma proteomics

Proteomic profiling of plasma samples was conducted using the SomaScan^®^ assay, a modified aptamer-based technology capable of quantifying plasma levels of ∼5000 proteins.^19^ Specificity and stability of the Assay has been extensively described.^20^ The total number of samples in each cohort available for proteomic analyses are 458, 298, and 104 baseline samples and an additional 491, 335, and 165 follow-up longitudinal samples in cohorts 1, 2 and 3, respectively, after removal of samples that did not pass assay QC criteria.

### Derivation and validation of Residual Cardiovascular Disease (RCV) model

Machine learning techniques were employed to develop the RCV model, resulting in a 27-protein model of composite cardiac event risk within four years (including myocardial infarction, heart failure hospitalizations, stroke or all-cause death). The model was developed and validated on SomaScan^®^ plasma proteomic data from 18,163 individuals in six different clinical studies. Prognostic performance of this test was assessed in multiple high-risk, multi-morbid patient populations, outperforming existing cardiovascular risk predictors. This model also demonstrated sensitivity to detecting changes to cardiovascular risk with approaching events and following interventions in the EXSCEL and DiRECT trials.^14^

### Statistical methods

The goals of the statistical analysis had multiple components, with the overarching purpose of understanding the prognostic performance of the RCV SomaSignal test from the first blood draw and comparing this with other laboratory and clinical measures.

Summary statistics (mean, standard deviation, range, percentages) were calculated on baseline demographics and lab measurements. The prognostic performance of the RCV model and competing metrics was assessed in two ways: 1) using regression models, which allowed us to characterize the effects of different covariates (e.g., RCV predictions, lab measurements) and determine which were statistically significant predictors of COVID-19 outcomes, and 2) using machine learning techniques and predictive metrics, which allowed to understand the predictive accuracy of the different covariates.

The two endpoints of interest were death (as a yes/no status variable) and the maximum severity the subject reached over the course of the study (as an ordinal variable). For the death endpoint, logistic regression models were used, and for the maximum severity endpoint, ordinal regression models were used. For application of machine learning techniques, each cohort was split into an 80/20 training and test set, with 5 repeats of 5-fold cross-validation performed on the training set and validation of the cross-validated model performed on the test set. Logistic regression classification models (for the death endpoint) were assessed using the area under the curve (AUC), sensitivity, and specificity. The ordinal regression models (for the maximum severity endpoint) were assessed using the root-mean square error (RMSE).

These analyses were performed to assess a number of components of the data. The first assessment was done on the RCV model predictions themselves in an effort to characterize how well the probabilities predict death and maximum severity from baseline. Following, this approach was used to examine the individual proteins in the RCV model, which allowed us to assess if a small subset of the proteins were driving the association between the RCV model predictions and COVID-19 outcomes. Lastly, this approach was used to quantify differences between the RCV model and various competing lab metrics as well as pre-existing heart conditions (this was done in the MGH cohort only, as it was the only cohort that had this clinical information). Only the baseline samples were used for this portion of the analyses.

Another goal of the analysis was to characterize the trajectory of the RCV model predictions over the course of the illness using the longitudinal samples. To this end, a repeated measures linear regression model was used, with RCV model predictions used as the endpoint (which were logit transformed to satisfy normality assumptions). In this model, the only predictor included was time, with time at the peak RCV predicted probabilities centered at 0 in order to align different patients’ natural histories, and time before peak probability as negative (with values farther away from zero being larger negative numbers) and time after peak probability as positive. Separate slopes were used to quantify the rate of RCV increase before peak severity and the rate of RCV decrease post-peak severity. This allowed us to predict a trajectory of RCV scores as well as determine if the rates of increase and decrease were statistically significant.

All calculations were performed in R v3.6.3.

## RESULTS

### Prognostic performance of 27-protein RCV model in predicting COVID-19 outcomes

Logistic regression models show a statistically significant increase in the risk of death as RCV predictions increase. Additionally, cross-validated models show AUC values of 82%, 79%, and 65% for the MSSM, MGH, and ISB test cohorts, respectively. See Table 2 for details.

In a similar fashion, boxplots of the baseline RCV predictions, stratified by maximum COVID-19 severity status, show a strong trend of increase in probability as the COVID-19 severity increases (see Figure 1). Additionally, ordinal logistic regression models show RCV to be a significant predictor of maximum severity (p < 0.01 for all cohorts). See Table 2 for details.

**Figure 1.**
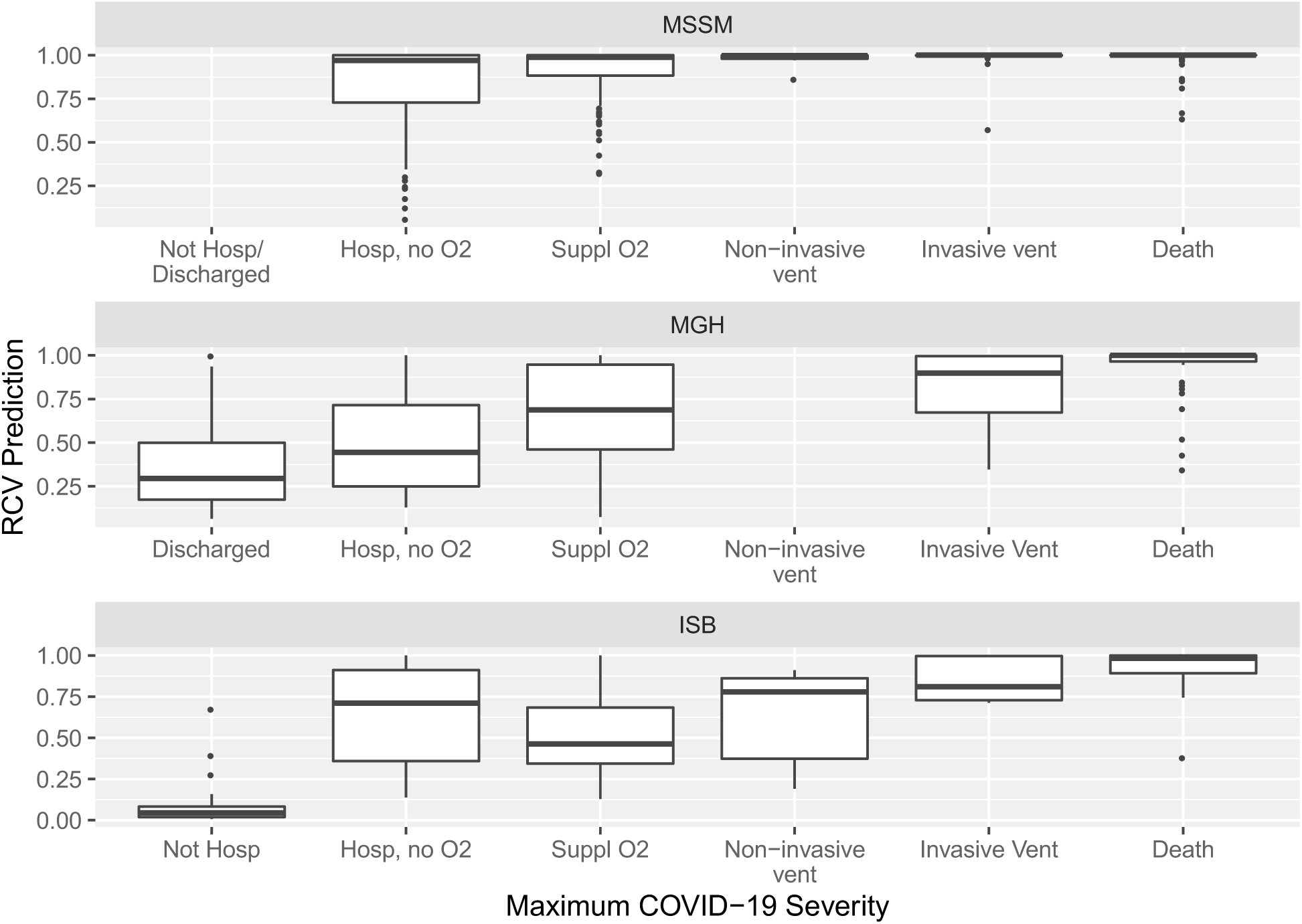
Boxplots of the baseline RCV predictions (y-axis), stratified by maximum COVID-19 severity status (x-axis). It can be observed that the probability of an event increases with severity, with the group of individuals who will die from COVID-19 having much higher probabilities than those who were discharged or not hospitalized. Hosp, hospitalized; Suppl, supplemental; vent, ventilation. MSSM, Mt Sinai School of Medicine; MGH, Massachusetts General Hospital; ISB, Institute for Systems Biology. Sample sizes for severity groupings in each cohort are shown in detail in Supplementary Table 1.

### Prognostic individual performance of the 27-proteins in the RCV model

In an effort to understand if a certain subset of the proteins in the RCV model had a greater association with COVID-19 outcomes, regression models and predictive model metrics (AUC for the death endpoint, RMSE for the maximum severity score) were calculated on a univariate level for each protein as it related to each endpoint. Additionally, the effect sizes from the regression coefficients were compared to each other by calculating the percent each estimated effect had out of the sum of all 27 absolute effect sizes. Protein measurements were log-10 transformed and centered/scaled before analysis so that estimated effect sizes were comparable.

Results are shown in Figure 2 (exact numbers are in Supplementary Table 3). It can be observed that, on a univariate level, protein effects range from 1% to 6.8% in percent effect size in prediction of death, which translates to a 120% to 380% increase in risk of death for every 1-unit increase in the standard deviation of the protein RFU measurement. It can also be observed that the predictive capabilities of each protein, on a univariate level, range from and AUC of 54% (marginally better than random chance) to 79%. Similarly, the percent estimated effect sizes range from 0.3% to 7.7% in prediction of the maximum severity score, with the biggest contributor translating to a 396% increase in severity for every 1-unit increase in the standard deviation of the protein RFU measurement.

**Table 3.** Predictive comparison (for death as an endpoint) between the RCV predictions and previous existing heart condition status (a binary variable) in the MGH cohort. The first row contains the results (balanced accuracy, specificity, and sensitivity) for the RCV model when the probability cut-off is chosen to maximize balanced accuracy, and the second row contains the results for a previous existing heart condition. The third row contains the results when the probability cut-off is chosen so that the specificity matches that of a previous existing heart condition.

| Predictor | Balanced Accuracy | Specificity | Sensitivity |
| --- | --- | --- | --- |
| RCV prediction<br>(cut-off is 98.67%, chosen to maximize balanced accuracy) | 0.78 | 0.84 | 0.73 |
| Previous existing heart condition | 0.63 | 0.88 | 0.39 |
| RCV risk, adjusted cut-off<br>(cut-off is 99.95%, chosen to match specificity of pre-existing heart condition) | 0.73 | 0.88 | 0.59 |

**Figure 2.**
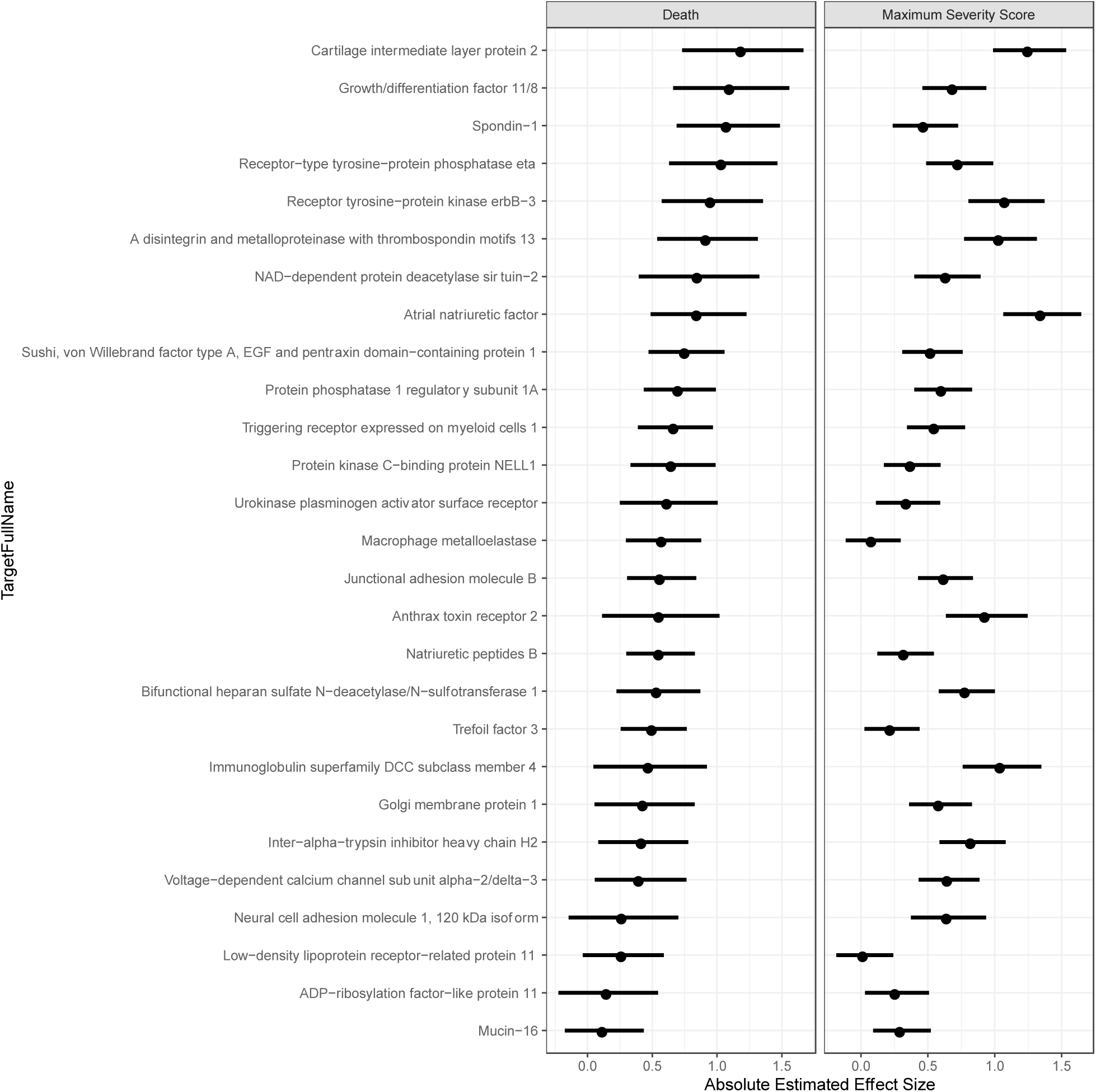
Forest plots for the absolute estimated effects of each individual protein in the RCV model, estimated on a univariate level for death and maximum severity score endpoints. The dot represents the absolute value of the effect size, and the lines represent the 95% confidence intervals of the effect, calculated using the absolute value.

### Comparison of RCV protein model to clinical variables and lab chemistry values

In assessing the performance of the cardiovascular protein model, another key component was the comparison of the performance of the model predictions compared to other standard lab metrics. To this end, using the MGH cohort, we compared the RCV model predictions to commonly screened biomarkers of cardiac injury, inflammation and immune response including, C-Reactive protein (CRP), D-Dimer, ferritin, fibrinogen, interleukin-6 (IL-6), NT-proBNP, procalcitonin, and high sensitivity cardiac Troponin (Hs-cTn). In this analysis, all measurements were centered and scaled so that the effect sizes could be comparable, with results expressed in standard deviation units. In addition to comparison to the lab metrics, we also compared the model to clinical information on whether the individual had a pre-existing heart condition (yes/no) so that we could understand if the RCV model was only explaining existing comorbidities or was providing additional risk information.

Results are shown in Figure 3 and full descriptive results outlined in Supplementary Table 4. It can be observed that the RCV model has the highest estimated effect size for both endpoints at 40% and 30% for death and maximum severity score, respectively. The next largest percent effect sizes are for D-Dimer (at 10.8%) for association with death and CRP (18.5%) for prediction of maximum severity scores. Additionally, the RCV model predictions have the highest AUC (83.5%) and lowest RMSE (1.3985), with the next highest AUC equal to 81.4% for Hs-cTn and the next lowest RMSE equal to 1.556 for D-Dimer. This provides strong evidence that the RCV model is a robust predictor of both death and maximum severity, particularly when compared to other standard lab metrics.

**Table 4.** The estimated trajectories for the RCV predictions for each cohort through the course of acute COVID-19 infection into convalescence (back-transformed from the logit scale). MSSM, Mt Sinai School of Medicine; MGH, Massachusetts General Hospital; ISB, Institute for Systems Biology.

| Covariate | MSSM | MGH | ISB |
| --- | --- | --- | --- |
| Average RCV score at peak | 99.996% (< 0.01) | 99.890% (< 0.01) | 61.893% (0.281) |
| Daily RCV increase approaching peak | 76% (< 0.01) | 191% (< 0.01) | 1.5% (0.037) |
| Daily RCV decrease post-peak | 60% (< 0.01) | 80% (< 0.01) | 4.8% (< 0.01) |

**Figure 3.**
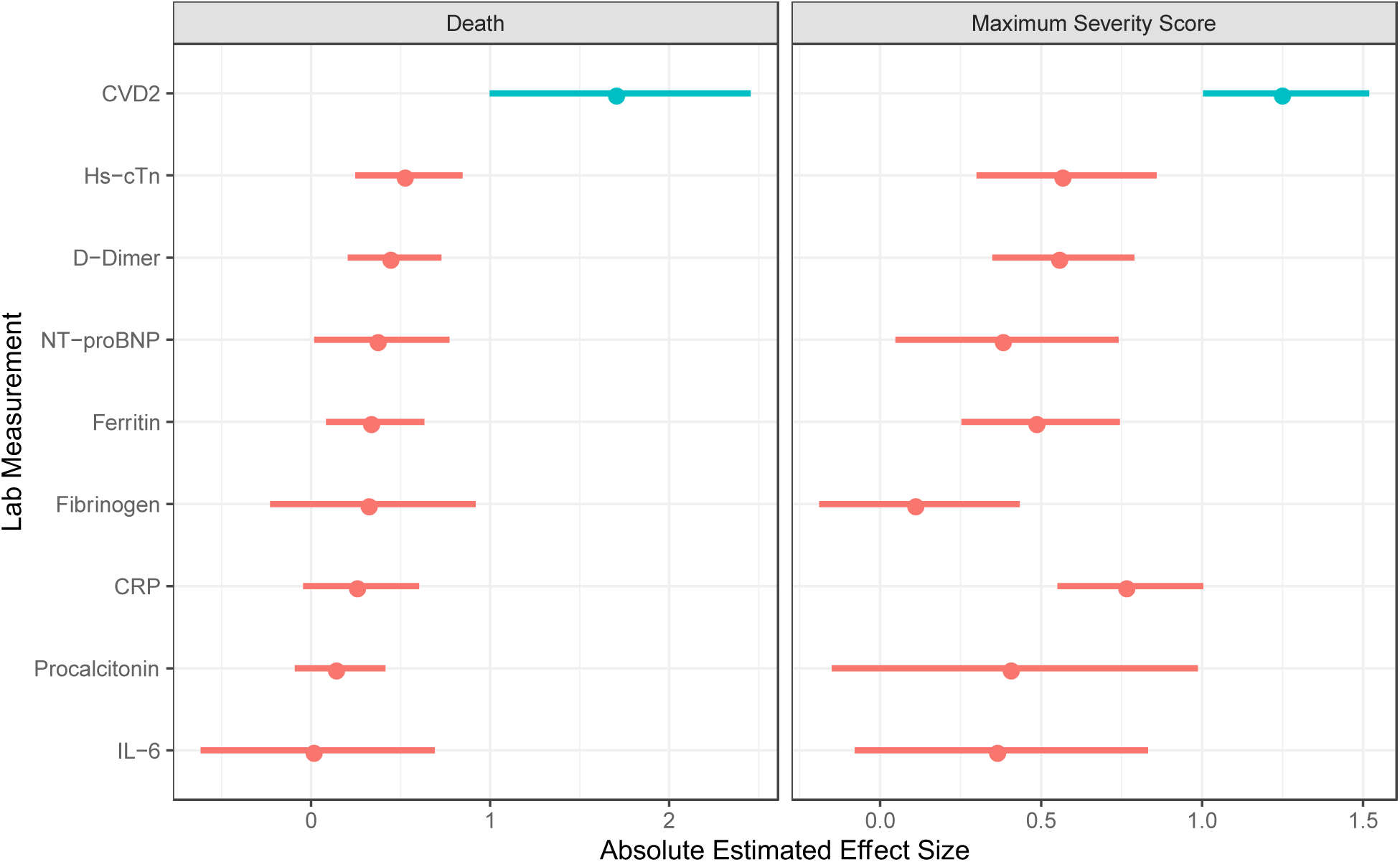
Forest plots of the absolute estimated effects of commonly used laboratory baseline measurements compared to the RCV model, estimated on a univariate level, in the prediction of death and maximum severity score. The dot represents the absolute value of the effect size, and the lines represent the 95% confidence intervals of the effect, calculated using the absolute value. ^*^Note: IL-6 and procalcitonin values were available for a subset of patients.

Table 3 shows results comparing the predictive accuracy of a known previous heart condition (a known major risk factor in COVID-19 severity) to the RCV predictions. This status is a binary yes/no variable (with “yes” indicating a subject died during the course of study observation). The RCV predictions are stratified by a selected cut-off point, with probabilities above the cut-off point designated as a “yes” prediction and those below labeled as “no.” When the cut-off probability is chosen such that balanced accuracy is maximized (cut-off equal to 98.67%), we observe that the RCV predictions are more accurate than a known heart condition (0.78 vs. 0.63). However, we see that previous heart condition has a higher specificity rate, which means that this metric is more accurate in identifying individuals who will not die from COVID-19. If the cut-off for the RCV probabilities is adjusted to 99.95%, then the RCV model specificity increases to match the heart condition specificity, and we see that the sensitivity (and therefore the balanced accuracy) is still higher for the RCV model. These results provide evidence that the RCV model is capturing addition risk outside of an already existing heart condition and is a flexible method for prediction of COVID-19 fatality.

### Sensitivity of the 27-protein RCV model to detect changes in cardiovascular risk across the course of COVID-19

It is possible that the RCV model predictions follow a trajectory that elucidates a mechanistic time-course of COVID-19 severity, independent of oxygen status or observed symptoms. To test this hypothesis, random effects models were calculated using the RCV model predictions as the response and time as a predictor. In this instance, time (in days) was calculated such that the peak RCV prediction occurred at time 0, time before the peak was negative, and time after the peak was positive. In the model, separate slopes were allowed for pre- and post-peak trajectories, and the RCV predictions were logit transformed to ensure back-transformed predictions between zero and one.

Results from the model can be seen in Table 4 and Figure 4. Most importantly, there were striking acute daily increases in predicted risks before the peak, and rapid decreases in scores after the peak which were are all statistically significant at the 0.05 level across all three cohorts, with rates of decrease similar to rates of increase. The predicted trajectories, along with the observed measurements, can be viewed in Figure 4.

**Figure 4.**
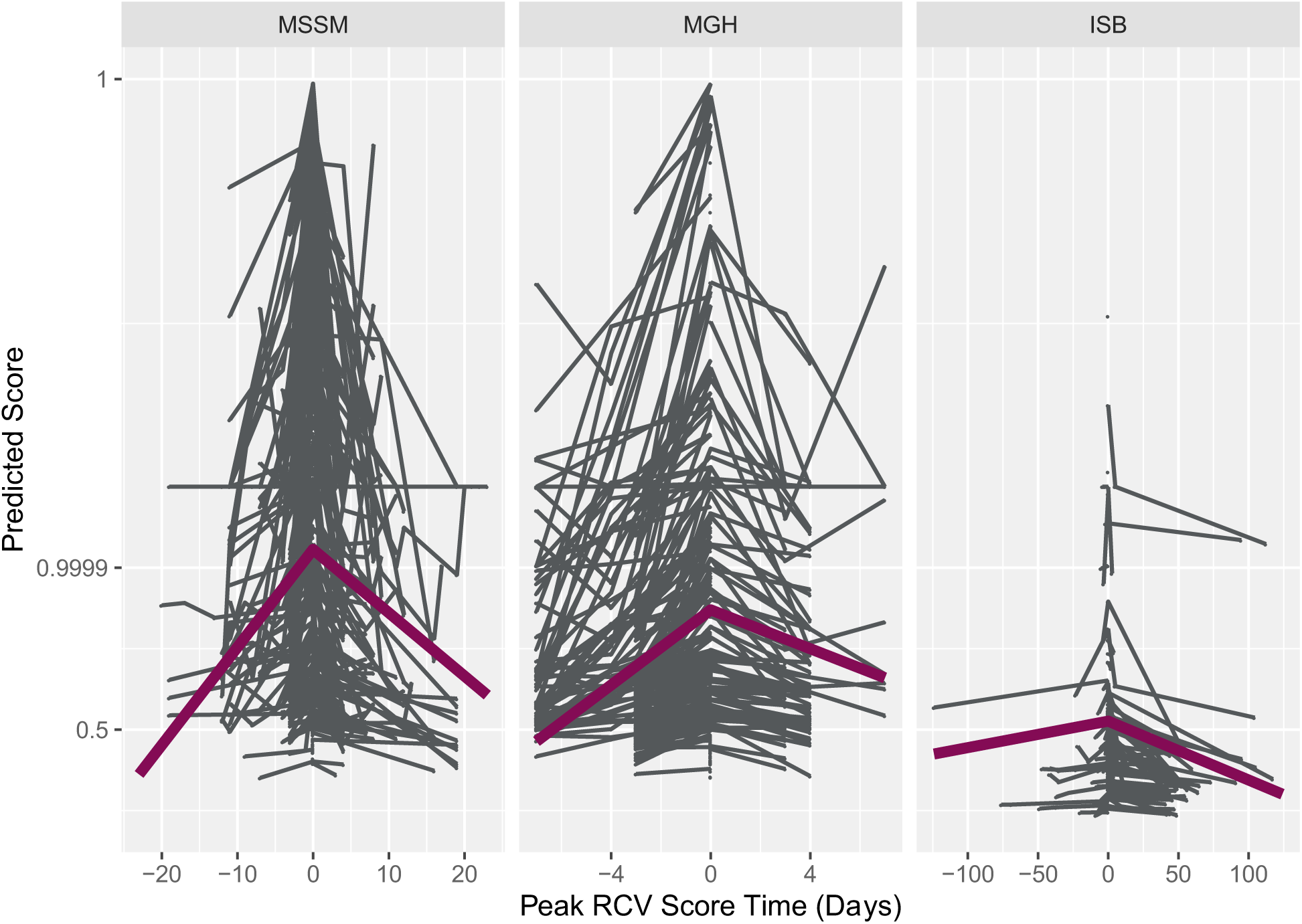
RCV trajectory for COVID-19 patients across the course of illness. The grey lines denote the observed RCV predictions for each subject, and the solid magenta line denotes the fitted line for the trajectories in each cohort. Time is along the x-axis (calculated such that zero is at the peak RCV prediction for each person), and the y-axis is the prediction. Note that the trajectory models were fit and are shown in the figure above using the logit transformation of the RCV prediction. However, the ticks on the y-axis note the corresponding probabilities to the logit value. MSSM, Mt Sinai School of Medicine; MGH, Massachusetts General Hospital; ISB, Institute for Systems Biology.

## DISCUSSION

COVID-19 is characterized by a heterogenous clinical course. While most patients experience only mild symptoms, many develop severe disease that is not fully explained by traditional risk factors or underlying comorbidities. Moreover, a significant proportion of patients harbor long-term health concerns post-infection, irrespective of initial disease severity. With no cure for COVID-19 in the near future and a lengthy roll-out period for newly developed vaccines, there is an urgent unmet need for reliable predictors of disease severity stratification as well as post-acute monitoring tools.

This study involved a 27-protein candidate surrogate endpoint, previously discovered using machine-learning applied to 5000 measured proteins in thousands of individuals with known outcomes. It was also previously validated at large scale in different chronic disease morbidities prior to the pandemic in people with COVID-19. In this study, we found that it predicted COVID-19 patients outcome severity and mortality risk when applied to samples at the time of presentation in both hospitalized and non-hospitalized individuals, with the median predicted risk in people who subsequently died across studies exceeding 95%. This suggests that COVID-19 is acting acutely upon the same biologic mechanisms that mediate chronic cardiovascular risk, and potentially that some of the same therapies might be applied to reduce that risk. Additionally, the longitudinal change from sequential samples in the same subjects sheds some light on the duration on the rate of risk ascendancy during symptomatic infection (predicted 1.0%-2.6% increase in absolute risk per day before the peak), and the rate of resolution in the survivors (predicted 0.3%-1% decrease in absolute risk per day post peak), and predicted risks were catastrophically high (above 50%) for a typical period of 8-12 days.

The 27 proteins in the RCV test represent cardiac function, metabolism, immune activation, inflammation, and fibrinolysis. Proposed mechanisms of cardiac events and cardiac damage observed in COVID-19 patients are not fully understood, but are thought to include systemic immune activation, direct impact of SARS-CoV-2 on the myocardium, or the consequence of ischemia/hypoxia/thrombosis.^21^ While these patients may have unknown subclinical cardiovascular disease risk prior to COVID-19, (accurately predicted by the 27-protein model^14^), the dramatic increases in risk predictions during infection suggest that prior disease is an incomplete explanation.

Additionally, given that all the proteins in the 27 protein model contributed to the prognostic prediction (each protein individually contributed 0.3-7.7% to the observed effect size), this suggests that the etiology of elevated RCV risk score in COVID-19 patients with severe outcomes is likely multifactorial and may explain why the RCV test offers greater predictive performance compared to traditional CV and immune biomarkers in the current study. As well as outperforming other blood-based biomarkers, our initial findings show that the RCV test may also outperform a number of other more complex, demographics-driven prognostic approaches recently published.^22,23^ Furthermore, since our model is protein-based and not dependent upon immutable factors, our analysis had the capability of extending beyond prognostication and allowed the natural history of biologic mechanisms associated with cardiovascular risk to be observed across the course of COVID-19 illness. We provide novel findings demonstrating the CV risk ascendancy and decline across acute COVID-19 infection into recovery. In this study we were not enabled to relate recovery RCV scores to subsequent cardiovascular event rates, but given that the 27-protein cardiovascular risk test has displayed prognostic sensitivity in other disease states,^14^ and was also prognostic during the ascendancy of infection, this deserves further follow-up as it may represent a scalable method of tracking residual risk during recovery.

Together, our results emphasize the importance of cardiovascular systems biology in COVID-19 outcomes and recovery. We present proteomic support for biological mechanisms involving cardiovascular dysfunction across the course of COVID-19 illness, which is in keeping with clinical observations that prior cardiovascular disease is both a causative risk factor, as well as an outcome in many patients with COVID-19,^10-12^ and up to 78% of COVID-19 survivors display cardiac involvement presenting long-lasting health concerns beyond the initial SARS-CoV-2 infection period.^24^

One limitation of the current study is that the criteria for hospitalization and diagnosis for COVID-19 are continuously evolving, as are the standard of care procedures for COVID-19 patients nationally and internationally. Therefore, the current findings require further replication in additional cohorts to ensure their validity. To address these limitations, we have utilized only patients with COVID-19 diagnosis confirmed by PCR methods. Moreover, our results hail from three independent studies spanning a collection timeframe of March-October. Another limitation of this study is that the prognostic performance of the residual CV risk model on COVID-19 outcomes may be improved by including other clinical variables that are believed to influence outcomes to SARS-CoV-2 infection, such as age, sex, BMI and laboratory blood chemistry values. However, we purposefully chose to not include those variables to a) allow greater sensitivity to change, and b) to favor the utility of the test in situations where extensive blood work, or chart review is not warranted i.e. in non-hospitalized settings.

Another limitation is that the SomaScan^®^ platform, upon which this test is based, takes ∼30 hours to deliver a result and is currently only available in a single central CAP/CLIA certified laboratory; near-patient use would require its translation onto another platform. This may mean that it is more useful during the recovery process than acutely, although this remains to be demonstrated.

We conclude that the same cardiovascular risk mechanisms active in chronic disease and captured by the 27-protein surrogate endpoint are also acutely enhanced during COVID-19 infection, that a period of catastrophic risk-elevation of 8-12 days ensues, followed by a resolution period that needs further characterization; this could be tracked by means of the 27-protein surrogate used in this study.

## Data Availability

The datasets analyzed during the current study are not publicly available due to United States Federal Health Insurance Portability and Accountability Act (HIPAA) compliance.

## Acknowledgements

The authors thank the SomaLogic Assay and Production Bioinformatics teams for sample assessment and the bioinformatics of quality control, respectively. The authors also thank M. Messenbaugh, E. King, S. Wilcox, and J. Williams for leading efforts in SomaLogic’s COVID-19 initiative.

