## Supplemental figures for "Application of a 27-protein candidate cardiovascular surrogate endpoint to track risk ascendancy and resolution in COVID-19"

### SUPPLEMENTARY FIGURES

#### A) MSSM

| Variable | | Total | Death | | Maximum COVID-19 severity | | | | |
|---|---|---|---|---|---|---|---|---|---|
| | | | No | Yes | Room Air | Supplemental O2 | Non-invasive Vent | Invasive Vent | Death |
| **Total** | | 458 (100) | 365 (80) | 93 (20) | 142 (31) | 150 (33) | 7 (2) | 20 (4) | 93 (20) |
| **Sex** | **Female** | 180 (39) | 138 (77) | 42 (23) | 58 (32) | 53 (29) | 5 (3) | 7 (4) | 42 (23) |
| | **Male** | 278 (61) | 227 (82) | 51 (18) | 84 (30) | 97 (35) | 2 (1) | 13 (5) | 51 (18) |
| **Ethnicity** | **Hispanic/Latino** | 186 (41) | 143 (77) | 43 (23) | 47 (25) | 63 (34) | 5 (3) | 11 (6) | 43 (23) |
| | **Not Hispanic/Latino** | 264 (58) | 214 (81) | 50 (19) | 91 (34) | 85 (32) | 2 (1) | 9 (3) | 50 (19) |
| | **Unknown** | 8 (2) | 8 (100) | 0 (0) | 4 (50) | 2 (25) | 0 (0) | 0 (0) | 0 (0) |
| **Race** | **American Indian/Alaska Native** | 8 (2) | 6 (75) | 2 (25) | 3 (38) | 3 (38) | 0 (0) | 0 (0) | 2 (25) |
| | **Asian** | 31 (7) | 28 (90) | 3 (10) | 8 (26) | 15 (48) | 1 (3) | 1 (3) | 3 (10) |
| | **Black or African American** | 109 (24) | 90 (83) | 19 (17) | 41 (38) | 35 (32) | 1 (1) | 1 (1) | 19 (17) |
| | **More Than One Race** | 24 (5) | 18 (75) | 6 (25) | 6 (25) | 8 (33) | 1 (4) | 1 (4) | 6 (25) |
| | **Native Hawaiian or Other Pacific Islander** | 2 (0) | 2 (100) | 0 (0) | 1 (50) | 1 (50) | 0 (0) | 0 (0) | 0 (0) |
| | **Unknown / Not Reported** | 119 (26) | 102 (86) | 17 (14) | 36 (30) | 44 (37) | 2 (2) | 9 (8) | 17 (14) |
| | **White** | 165 (36) | 119 (72) | 46 (28) | 47 (28) | 44 (27) | 2 (1) | 8 (5) | 46 (28) |
| **BMI** | **Underweight** | 22 (5) | 16 (73) | 6 (27) | 9 (41) | 4 (18) | 0 (0) | 1 (5) | 6 (27) |
| | **Normal** | 123 (27) | 98 (80) | 25 (20) | 40 (33) | 36 (29) | 2 (2) | 9 (7) | 25 (20) |

| | | | | | | | | | |
|---|---|---|---|---|---|---|---|---|---|
| | **Overweight** | 145 (32) | 113 (78) | 32 (22) | 43 (30) | 52 (36) | 1 (1) | 3 (2) | 32 (22) |
| | **Obese** | 128 (28) | 105 (82) | 23 (18) | 40 (31) | 41 (32) | 2 (2) | 7 (5) | 23 (18) |
| | **Severely Obese** | 32 (7) | 26 (81) | 6 (19) | 9 (28) | 14 (44) | 1 (3) | 0 (0) | 6 (19) |
| | **Unknown** | 8 (2) | 7 (88) | 1 (12) | 1 (12) | 3 (38) | 1 (12) | 0 (0) | 1 (12) |
| **Asthma** | **No** | 370 (94) | 291 (79) | 79 (21) | 132 (36) | 135 (36) | 5 (1) | 19 (5) | 79 (21) |
| | **Yes** | 25 (6) | 19 (76) | 6 (24) | 7 (28) | 11 (44) | 1 (4) | 0 (0) | 6 (24) |
| **COPD** | **No** | 365 (92) | 290 (79) | 75 (21) | 131 (36) | 136 (37) | 5 (1) | 18 (5) | 75 (21) |
| | **Yes** | 30 (8) | 20 (67) | 10 (33) | 8 (27) | 10 (33) | 1 (3) | 1 (3) | 10 (33) |
| **HTN** | **No** | 221 (56) | 177 (80) | 44 (20) | 70 (32) | 94 (43) | 2 (1) | 11 (5) | 44 (20) |
| | **Yes** | 174 (44) | 133 (76) | 41 (24) | 69 (40) | 52 (30) | 4 (2) | 8 (5) | 41 (24) |
| **Diabetes** | **No** | 283 (72) | 226 (80) | 57 (20) | 97 (34) | 110 (39) | 3 (1) | 16 (6) | 57 (20) |
| | **Yes** | 112 (28) | 84 (75) | 28 (25) | 42 (38) | 36 (32) | 3 (3) | 3 (3) | 28 (25) |
| **CKD** | **No** | 332 (84) | 267 (80) | 65 (20) | 113 (34) | 130 (39) | 5 (2) | 19 (6) | 65 (20) |
| | **Yes** | 63 (16) | 43 (68) | 20 (32) | 26 (41) | 16 (25) | 1 (2) | 0 (0) | 20 (32) |
| **CAD** | **No** | 334 (85) | 269 (81) | 65 (19) | 111 (33) | 135 (40) | 6 (2) | 17 (5) | 65 (19) |
| | **Yes** | 61 (15) | 41 (67) | 20 (33) | 28 (46) | 11 (18) | 0 (0) | 2 (3) | 20 (33) |
| **Heart Failure** | **No** | 346 (88) | 274 (79) | 72 (21) | 114 (33) | 138 (40) | 4 (1) | 18 (5) | 72 (21) |
| | **Yes** | 49 (12) | 36 (73) | 13 (27) | 25 (51) | 8 (16) | 2 (4) | 1 (2) | 13 (27) |
| **Smoking Status** | **NEVER** | 183 (46) | 151 (83) | 32 (17) | 67 (37) | 70 (38) | 3 (2) | 11 (6) | 32 (17) |
| | **NOT ASKED** | 2 (1) | 2 (100) | 0 (0) | 0 (0) | 2 (100) | 0 (0) | 0 (0) | 0 (0) |
| | **QUIT** | 121 (31) | 91 (75) | 30 (25) | 52 (43) | 34 (28) | 2 (2) | 3 (2) | 30 (25) |
| | **YES** | 28 (7) | 19 (68) | 9 (32) | 9 (32) | 7 (25) | 1 (4) | 2 (7) | 9 (32) |
| | **Unknown** | 61 (15) | 47 (77) | 14 (23) | 11 (18) | 33 (54) | 0 (0) | 3 (5) | 14 (23) |
| **Cancer** | **No** | 355 (90) | 277 (78) | 78 (22) | 121 (34) | 133 (37) | 6 (2) | 17 (5) | 78 (22) |
| | **Yes** | 40 (10) | 33 (82) | 7 (18) | 18 (45) | 13 (32) | 0 (0) | 2 (5) | 7 (18) |
| **Age** | **Mean** | 63.28 (16.08) | 61.16 (16.19) | 71.59 (12.63) | 63.07 (16.68) | 60.25 (15.82) | 62.71 (18.36) | 58.25 (15.77) | 71.59 (12.63) |
| | **Range** | (20, 90) | (20, 90) | (46, 90) | (20, 90) | (20, 90) | (41, 90) | (24, 84) | (46, 90) |

**B) MGH**

| Variable | | Total | Death | | Maximum COVID-19 severity | | | | |
|---|---|---|---|---|---|---|---|---|---|
| | | | No | Yes | Discharged | No O2 | Supplemental O2 | Invasive vent | Death |
| **Total** | | 298 (100) | 257 (86) | 41 (14) | 28 (9) | 35 (12) | 130 (44) | 64 (21) | 41 (14) |
| **Sex** | **Female** | 142 (48) | 124 (87) | 18 (13) | 18 (13) | 17 (12) | 62 (44) | 27 (19) | 18 (13) |
| | **Male** | 156 (52) | 133 (85) | 23 (15) | 10 (6) | 18 (12) | 68 (44) | 37 (24) | 23 (15) |
| **Ethnicity** | **Hispanic/Latino** | 161 (54) | 150 (93) | 11 (7) | 12 (7) | 24 (15) | 71 (44) | 43 (27) | 11 (7) |
| | **Not Hispanic/Latino** | 137 (46) | 107 (78) | 30 (22) | 16 (12) | 11 (8) | 59 (43) | 21 (15) | 30 (22) |
| **Race** | **White** | 151 (51) | 123 (81) | 28 (19) | 15 (10) | 13 (9) | 75 (50) | 20 (13) | 28 (19) |
| | **Black** | 28 (9) | 24 (86) | 4 (14) | 1 (4) | 5 (18) | 10 (36) | 8 (29) | 4 (14) |
| | **Asian** | 11 (4) | 9 (82) | 2 (18) | 3 (27) | NA (NA) | 4 (36) | 2 (18) | 2 (18) |
| | **Other** | 108 (36) | 101 (94) | 7 (6) | 9 (8) | 17 (16) | 41 (38) | 34 (31) | 7 (6) |
| **BMI** | **Underweight** | 2 (1) | 2 (100) | NA (NA) | NA (NA) | NA (NA) | 2 (100) | NA (NA) | NA (NA) |
| | **Normal** | 42 (14) | 35 (83) | 7 (17) | 2 (5) | 8 (19) | 15 (36) | 10 (24) | 7 (17) |
| | **Overweight** | 107 (36) | 89 (83) | 18 (17) | 6 (6) | 15 (14) | 43 (40) | 25 (23) | 18 (17) |
| | **Obese** | 93 (31) | 85 (91) | 8 (9) | 7 (8) | 8 (9) | 50 (54) | 20 (22) | 8 (9) |
| | **Severely Obese** | 34 (11) | 29 (85) | 5 (15) | 1 (3) | 2 (6) | 18 (53) | 8 (24) | 5 (15) |
| | **Unknown** | 20 (7) | 17 (85) | 3 (15) | 12 (60) | 2 (10) | 2 (10) | 1 (5) | 3 (15) |
| **Asthma** | **No** | 252 (85) | 216 (86) | 36 (14) | 25 (10) | 27 (11) | 106 (42) | 58 (23) | 36 (14) |
| | **Yes** | 46 (15) | 41 (89) | 5 (11) | 3 (7) | 8 (17) | 24 (52) | 6 (13) | 5 (11) |
| **COPD** | **No** | 273 (92) | 238 (87) | 35 (13) | 27 (10) | 34 (12) | 115 (42) | 62 (23) | 35 (13) |
| | **Yes** | 25 (8) | 19 (76) | 6 (24) | 1 (4) | 1 (4) | 15 (60) | 2 (8) | 6 (24) |
| **HTN** | **No** | 157 (53) | 146 (93) | 11 (7) | 21 (13) | 22 (14) | 72 (46) | 31 (20) | 11 (7) |
| | **Yes** | 141 (47) | 111 (79) | 30 (21) | 7 (5) | 13 (9) | 58 (41) | 33 (23) | 30 (21) |
| **DIABETES** | **No** | 191 (64) | 166 (87) | 25 (13) | 22 (12) | 24 (13) | 89 (47) | 31 (16) | 25 (13) |
| | **Yes** | 107 (36) | 91 (85) | 16 (15) | 6 (6) | 11 (10) | 41 (38) | 33 (31) | 16 (15) |
| **CKD** | **No** | 259 (87) | 232 (90) | 27 (10) | 27 (10) | 32 (12) | 119 (46) | 54 (21) | 27 (10) |

| | | | | | | | | | |
|---|---|---|---|---|---|---|---|---|---|
| | **Yes** | 39 (13) | 25 (64) | 14 (36) | 1 (3) | 3 (8) | 11 (28) | 10 (26) | 14 (36) |
| **CAD** | **No** | 271 (91) | 239 (88) | 32 (12) | 27 (10) | 32 (12) | 119 (44) | 61 (23) | 32 (12) |
| | **Yes** | 27 (9) | 18 (67) | 9 (33) | 1 (4) | 3 (11) | 11 (41) | 3 (11) | 9 (33) |
| **Heart Failure** | **No** | 268 (90) | 237 (88) | 31 (12) | 28 (10) | 34 (13) | 116 (43) | 59 (22) | 31 (12) |
| | **Yes** | 30 (10) | 20 (67) | 10 (33) | NA (NA) | 1 (3) | 14 (47) | 5 (17) | 10 (33) |
| **Smoking status** | **Never or Unknown** | 212 (71) | 189 (89) | 23 (11) | 24 (11) | 28 (13) | 92 (43) | 45 (21) | 23 (11) |
| | **Current** | 75 (25) | 59 (79) | 16 (21) | 4 (5) | 4 (5) | 35 (47) | 16 (21) | 16 (21) |
| | **Prior** | 11 (4) | 9 (82) | 2 (18) | NA (NA) | 3 (27) | 3 (27) | 3 (27) | 2 (18) |
| **Cancer** | **No** | 284 (95) | 247 (87) | 37 (13) | 28 (10) | 33 (12) | 124 (44) | 62 (22) | 37 (13) |
| | **Yes** | 14 (5) | 10 (71) | 4 (29) | NA (NA) | 2 (14) | 6 (43) | 2 (14) | 4 (29) |
| **Age** | **mean** | 59.6 (18.69) | 56.44 (17.72) | 79.39 (10.97) | 79.39 (10.97) | 54.71 (17.23) | 42.86 (15.72) | 59.69 (14.25) | 58.23 (18.56) |
| | **range** | (25, 95) | (25, 95) | (45, 95) | (45, 95) | (25, 95) | (25, 85) | (25, 85) | (25, 95) |

##### C) ISB

| Variable | | Total | Death | | Maximum COVID-19 severity | | | | | |
|---|---|---|---|---|---|---|---|---|---|---|
| | | | No | Yes | Not hospitalized | No O2 | Supp O2 | Non-invasive Vent | Invasive Vent | Death |
| **Total** | | 104 (100) | 96 (92) | 8 (8) | 36 (35) | 8 (8) | 14 (13) | 7 (7) | 7 (7) | 8 (8) |
| **Sex** | **Female** | 62 (60) | 60 (97) | 2 (3) | 30 (48) | 4 (6) | 8 (13) | 2 (3) | 2 (3) | 2 (3) |
| | **Male** | 42 (40) | 36 (86) | 6 (14) | 6 (14) | 4 (10) | 6 (14) | 5 (12) | 5 (12) | 6 (14) |
| **Ethnicity** | **Hispanic/Latino** | 12 (12) | 12 (100) | 0 (0) | 3 (25) | 0 (0) | 1 (8) | 2 (17) | 2 (17) | 0 (0) |
| | **Not Hispanic/Latino** | 88 (85) | 80 (91) | 8 (9) | 30 (34) | 8 (9) | 13 (15) | 5 (6) | 4 (5) | 8 (9) |
| | **Unknown** | 4 (4) | 4 (100) | 0 (0) | 3 (75) | 0 (0) | 0 (0) | 0 (0) | 1 (25) | 0 (0) |
| **Race** | **American Indian/Alaska Native** | 1 (1) | 1 (100) | 0 (0) | 0 (0) | 0 (0) | 1 (100) | 0 (0) | 0 (0) | 0 (0) |
| | **Asian** | 15 (14) | 15 (100) | 0 (0) | 4 (27) | 1 (7) | 4 (27) | 1 (7) | 0 (0) | 0 (0) |
| | **Black or African American** | 7 (7) | 6 (86) | 1 (14) | 1 (14) | 2 (29) | 0 (0) | 2 (29) | 0 (0) | 1 (14) |
| | **More Than One Race** | 1 (1) | 1 (100) | 0 (0) | 1 (100) | 0 (0) | 0 (0) | 0 (0) | 0 (0) | 0 (0) |
| | **Native Hawaiian or Other Pacific Islander** | 3 (3) | 3 (100) | 0 (0) | 1 (33) | 1 (33) | 1 (33) | 0 (0) | 0 (0) | 0 (0) |
| | **Unknown / Not Reported** | 14 (13) | 14 (100) | 0 (0) | 4 (29) | 0 (0) | 2 (14) | 2 (14) | 2 (14) | 0 (0) |
| | **White** | 63 (61) | 56 (89) | 7 (11) | 25 (40) | 4 (6) | 6 (10) | 2 (3) | 5 (8) | 7 (11) |
| **BMI** | **Underweight** | 3 (3) | 2 (67) | 1 (33) | 0 (0) | 0 (0) | 0 (0) | 0 (0) | 0 (0) | 1 (33) |
| | **Normal** | 16 (15) | 13 (81) | 3 (19) | 0 (0) | 3 (19) | 5 (31) | 1 (6) | 1 (6) | 3 (19) |
| | **Overweight** | 23 (22) | 19 (83) | 4 (17) | 1 (4) | 0 (0) | 3 (13) | 3 (13) | 3 (13) | 4 (17) |
| | **Obese** | 21 (20) | 21 (100) | 0 (0) | 1 (5) | 5 (24) | 4 (19) | 3 (14) | 2 (10) | 0 (0) |
| | **Severely Obese** | 5 (5) | 5 (100) | 0 (0) | 0 (0) | 0 (0) | 2 (40) | 0 (0) | 1 (20) | 0 (0) |
| | **Unknown** | 36 (35) | 36 (100) | 0 (0) | 34 (94) | 0 (0) | 0 (0) | 0 (0) | 0 (0) | 0 (0) |

| | | | | | | | | | | |
|---|---|---|---|---|---|---|---|---|---|---|
| **Asthma** | **No** | 88 (85) | 80 (91) | 8 (9) | 26 (30) | 8 (9) | 12 (14) | 6 (7) | 7 (8) | 8 (9) |
| | **Yes** | 16 (15) | 16 (100) | 0 (0) | 10 (62) | 0 (0) | 2 (12) | 1 (6) | 0 (0) | 0 (0) |
| **COPD** | **No** | 97 (93) | 90 (93) | 7 (7) | 35 (36) | 8 (8) | 14 (14) | 5 (5) | 7 (7) | 7 (7) |
| | **Yes** | 7 (7) | 6 (86) | 1 (14) | 1 (14) | 0 (0) | 0 (0) | 2 (29) | 0 (0) | 1 (14) |
| **HTN** | **No** | 60 (58) | 55 (92) | 5 (8) | 29 (48) | 4 (7) | 8 (13) | 3 (5) | 4 (7) | 5 (8) |
| | **Yes** | 44 (42) | 41 (93) | 3 (7) | 7 (16) | 4 (9) | 6 (14) | 4 (9) | 3 (7) | 3 (7) |
| **Diabetes** | **No** | 81 (78) | 73 (90) | 8 (10) | 33 (41) | 7 (9) | 11 (14) | 5 (6) | 5 (6) | 8 (10) |
| | **Other** | 2 (2) | 2 (100) | 0 (0) | 2 (100) | 0 (0) | 0 (0) | 0 (0) | 0 (0) | 0 (0) |
| | **T1DM** | 2 (2) | 2 (100) | 0 (0) | 1 (50) | 1 (50) | 0 (0) | 0 (0) | 0 (0) | 0 (0) |
| | **T2DM** | 19 (18) | 19 (100) | 0 (0) | 0 (0) | 0 (0) | 3 (16) | 2 (11) | 2 (11) | 0 (0) |
| **CKD** | **No** | 97 (93) | 90 (93) | 7 (7) | 36 (37) | 7 (7) | 13 (13) | 6 (6) | 6 (6) | 7 (7) |
| | **Yes** | 7 (7) | 6 (86) | 1 (14) | 0 (0) | 1 (14) | 1 (14) | 1 (14) | 1 (14) | 1 (14) |
| **CAD** | **No** | 97 (93) | 92 (95) | 5 (5) | 35 (36) | 7 (7) | 12 (12) | 7 (7) | 7 (7) | 5 (5) |
| | **Yes** | 7 (7) | 4 (57) | 3 (43) | 1 (14) | 1 (14) | 2 (29) | 0 (0) | 0 (0) | 3 (43) |
| **Heart Failure** | **No** | 98 (94) | 91 (93) | 7 (7) | 36 (37) | 8 (8) | 11 (11) | 6 (6) | 7 (7) | 7 (7) |
| | **Yes** | 6 (6) | 5 (83) | 1 (17) | 0 (0) | 0 (0) | 3 (50) | 1 (17) | 0 (0) | 1 (17) |
| **Smoking Status** | **Current** | 4 (4) | 4 (100) | 0 (0) | 2 (50) | 0 (0) | 0 (0) | 1 (25) | 0 (0) | 0 (0) |
| | **Former** | 35 (34) | 30 (86) | 5 (14) | 11 (31) | 1 (3) | 2 (6) | 3 (9) | 3 (9) | 5 (14) |
| | **Never** | 59 (57) | 56 (95) | 3 (5) | 21 (36) | 7 (12) | 11 (19) | 3 (5) | 2 (3) | 3 (5) |
| | **Unknown** | 6 (6) | 6 (100) | 0 (0) | 2 (33) | 0 (0) | 1 (17) | 0 (0) | 2 (33) | 0 (0) |
| **Cancer** | **No** | 90 (87) | 86 (96) | 4 (4) | 31 (34) | 6 (7) | 14 (16) | 7 (8) | 7 (8) | 4 (4) |
| | **Yes** | 14 (13) | 10 (71) | 4 (29) | 5 (36) | 2 (14) | 0 (0) | 0 (0) | 0 (0) | 4 (29) |
| **Age** | **mean** | 57.075 (19.16) | 55.29 (18.77) | 78.38 (7.69) | 45.42 (15.98) | 64.38 (12.07) | 54.07 (14.87) | 65 (5.92) | 61.29 (20.16) | 78.38 (7.69) |
| | **range** | (18, 89) | (18, 89) | (67, 89) | (19, 81) | (40, 77) | (26, 80) | (56, 75) | (33, 86) | (67, 89) |

**Supplementary Table 1.** Summary demographics of COVID-19 patients by outcome in A) MSSM, B) MGH and C) ISB cohorts. Unless otherwise notes vales represent n (%). Abbreviations: BMI, Body Mass index; CAD, Coronary Artery Disease; CKD, Chronic Kidney Disease; COPD, Chronic Obstructive Pulmonary Disease; HTN, Hypertension; ISB, Institute for Systems Biology; MGH, Massachusetts General Hospital; MSSM, Mt Sinai School of Medicine. Note. Severity scores/clinical needed to generate scores were missing from a subset of patients.

| Cohort | Severity Rank | Clinical description |
|---|---|---|
| **MSSM** | 1 | Hospitalized,<br>no oxygen support |
| | 2 | Hospitalized, Supplemental oxygen support |
| | 3 | Hospitalized, non-invasive ventilation |
| | 4 | Hospitalized, invasive ventilation |
| | 5 | In-hospital death |
| **MGH** | 1 | Discharged from Emergency Department |
| | 2 | Hospitalized, no oxygen support |
| | 3 | Hospitalized, Supplemental oxygen support |
| | 4 | Hospitalized, invasive ventilation |
| | 5 | Death during follow up |
| **ISB** | 1 | Not hospitalized, ambulatory with no limitation of activities |
| | | Not hospitalized, ambulatory with limitation of activities |
| | 2 | Hospitalized, no oxygen support |
| | 3 | Hospitalized, oxygen by mask or nasal prongs |
| | 4 | Hospitalized, non-invasive ventilation |
| | 5 | Hospitalized, invasive ventilation |
| | | Hospitalized, invasive ventilation+organ support |
| | 6 | Death during follow up |

**Supplemental Table 2.** Clinical definitions of ordinal COVID-19 severity gradings used in each cohort in the current study. MSSM, Mt Sinai School of Medicine; MGH, Massachusetts General Hospital; ISB, Institute for Systems Biology.

| Target | Death | | | Maximum Severity | | |
|---|---|---|---|---|---|---|
| | Estimated effect | Percent of estimated effect | AUC** | Estimated effect | Percent of estimated effect | RMSE** |
| Cartilage intermediate layer protein 2 | -1.199 | 0.069 | 0.765 | -1.261 | 0.073 | 1.495 |
| Growth/differentiation factor 11/8 | -1.11 | 0.064 | 0.751 | -0.698 | 0.041 | 1.594 |
| Spondin-1 | 1.087 | 0.062 | 0.757 | 0.481 | 0.028 | 1.597 |
| Receptor-type tyrosine-protein phosphatase eta | -1.048 | 0.06 | 0.757 | -0.738 | 0.043 | 1.6 |
| Receptor tyrosine-protein kinase erbB-3 | -0.964 | 0.055 | 0.758 | -1.087 | 0.063 | 1.514 |
| A disintegrin and metalloproteinase with thrombospondin motifs 13 | -0.927 | 0.053 | 0.729 | -1.043 | 0.061 | 1.587 |
| NAD-dependent protein deacetylase sirtuin-2 | -0.861 | 0.049 | 0.703 | -0.647 | 0.038 | 1.606 |
| Atrial natriuretic factor | 0.858 | 0.049 | 0.723 | 1.356 | 0.079 | 1.512 |
| Sushi, von Willebrand factor type A, EGF and pentraxin domain-containing protein 1 | 0.764 | 0.044 | 0.775 | 0.534 | 0.031 | 1.642 |
| Protein phosphatase 1 regulatory subunit 1A | 0.712 | 0.041 | 0.733 | 0.615 | 0.036 | 1.589 |
| Triggering receptor expressed on myeloid cells 1 | 0.679 | 0.039 | 0.704 | 0.561 | 0.033 | 1.595 |
| Protein kinase C-binding protein NELL1 | -0.66 | 0.038 | 0.694 | -0.383 | 0.022 | 1.591 |
| Urokinase plasminogen activator surface receptor | 0.627 | 0.036 | 0.664 | 0.352 | 0.021 | 1.619 |
| Macrophage metalloelastase | 0.586 | 0.034 | 0.691 | 0.091 | 0.005 | 1.61 |
| Junctional adhesion molecule B | 0.572 | 0.033 | 0.742 | 0.632 | 0.037 | 1.635 |
| Anthrax toxin receptor 2 | -0.566 | 0.032 | 0.579 | -0.94 | 0.055 | 1.565 |
| Natriuretic peptides B | 0.564 | 0.032 | 0.7 | 0.333 | 0.019 | 1.613 |
| Bifunctional heparan sulfate N-deacetylase/N-sulfotransferase 1 | -0.547 | 0.031 | 0.668 | -0.791 | 0.046 | 1.587 |

| | | | | | | |
|---|---|---|---|---|---|---|
| Trefoil factor 3 | 0.511 | 0.029 | 0.735 | 0.232 | 0.013 | 1.616 |
| Immunoglobulin superfamily DCC subclass member 4 | -0.483 | 0.028 | 0.646 | -1.054 | 0.061 | 1.604 |
| Golgi membrane protein 1 | 0.441 | 0.025 | 0.638 | 0.595 | 0.035 | 1.642 |
| Inter-alpha-trypsin inhibitor heavy chain H2 | -0.43 | 0.025 | 0.63 | -0.834 | 0.049 | 1.632 |
| Voltage-dependent calcium channel subunit alpha-2/delta-3 | -0.41 | 0.023 | 0.61 | -0.658 | 0.038 | 1.618 |
| Neural cell adhesion molecule 1, 120 kDa isoform | -0.277 | 0.016 | 0.546 | -0.654 | 0.038 | 1.613 |
| Low-density lipoprotein receptor-related protein 11 | 0.275 | 0.016 | 0.574 | -0.029 | 0.002 | 1.61 |
| ADP-ribosylation factor-like protein 11 | -0.159 | 0.009 | 0.583 | -0.269 | 0.016 | 1.61 |
| Mucin-16 | 0.129 | 0.007 | 0.549 | -0.306 | 0.018 | 1.622 |

**Supplementary Table 3**. Univariate results and predictive metrics for each individual protein in the RCV model for COVID-related death and maximum severity score in the MGH cohort. * Not statistically significant at the 0.05 level. ** Not cross-validated.

| | Death | | | | Maximum severity score | |
|---|---|---|---|---|---|---|
| Lab measurement | Percent of total estimated effect size | Specificity | Sensitivity | AUC | Percent of total estimated effect | RMSE |
| RCV prediction | 0.400 | 0.837 | 0.732 | 0.835 | 0.252 | 1.385 |
| Hs-cTn | 0.127 | 0.785 | 0.769 | 0.814 | 0.116 | 1.578 |
| D-Dimer | 0.108 | 0.734 | 0.641 | 0.697 | 0.114 | 1.556 |
| NT-proBNP | 0.091* | 0.695 | 0.714 | 0.727 | 0.079 | 1.769 |
| Ferritin | 0.083 | 0.89 | 0.282 | 0.578 | 0.100 | 1.569 |
| Fibrinogen | 0.080* | 0.594 | 0.688 | 0.606 | 0.024* | 1.584 |
| CRP | 0.065* | 0.478 | 0.795 | 0.616 | 0.156 | 1.563 |
| Procalcitonin | 0.038* | 0.798 | 0.513 | 0.682 | 0.084* | 1.475 |
| IL-6 | 0.009* | 0.786 | 0.667 | 0.675 | 0.075* | 1.494 |

**Supplementary Table 4**. Predictive metrics for each individual common laboratory measurement obtained at baseline compared to the predictive performance of RCV model for COVID-related death and maximum severity score in the MGH cohort.CRP, C-Reactive Protein; IL-6, Interleukin-6; NT-proBNP, N-terminal pro-BNP; Hs-cTn, High sensitivity cardiac troponin. IL-6 and procalcitonin values were available for a subset of patients.* Not significant at the 0.05 level.
